# *IFI27* transcription is an early predictor for COVID-19 outcomes; a multi-cohort observational study

**DOI:** 10.1101/2021.10.29.21265555

**Authors:** Maryam Shojaei, Amir Shamshirian, James Monkman, Laura Grice, Minh Tran, Chin Wee Tan, Gustavo Rodrigues Rossi, Timothy R. McCulloch, Marek Nalos, Keng Yih Chew, Yanshan Zhu, Yao Xia, Timothy J. Wells, Alexandra Cristina Senegaglia, Carmen Lúcia Kuniyoshi Rebelatto, Claudio Luciano Franck, Anna Flavia Ribeiro dos Santos, Lucia de Noronha, Sepideh Motamen, Reza Valadan, Omolbanin Amjadi, Rajan Gogna, Esha Madan, Reza Alizadeh-Navaei, Liliana Lamperti, Felipe Zuñiga, Estefania Nova-Lamperti, Gonzalo Labarca, Ben Knippenberg, Velma Herwanto, Ya Wang, Amy Phu, Tracy Chew, Timothy Kwan, Karan Kim, Sally Teoh, Tiana M Pelaia, Win Sen Kuan, Yvette Jee, Jon Iredell, Ken O’Byrne, John F. Fraser, Melissa J. Davis, Gabrielle Belz, Majid Warkiani, Carlos Salomon Gallo, Fernando Souza-Fonseca-Guimaraes, Quan Nguyen, Anthony Mclean, Arutha Kulasinghe, Kirsty R. Short, Benjamin Tang

**Author notes:** Corresponding Authors: Dr Arutha Kulasinghe, The University of Queensland Diamantina Institute, The University of Queensland, 37 Kent Street, Woolloongabba, Queensland 4102, Australia, Dr Kirsty Short, School of Chemistry and Molecular Biology, Building 76, The University of Queensland, Brisbane, Queensland 4072, Australia, Dr Maryam Shojaei, Centre for Immunology and Allergy Research, Westmead Institute for Medical Research, Sydney, NSW, Australia. co-senior authors (Arutha Kulasinghe, Kirsty R. Short, Benjamin Tang).

## Abstract

**Background:** Robust biomarkers that predict disease outcomes amongst COVID-19 patients are necessary for both patient triage and resource prioritisation. Numerous candidate biomarkers have been proposed for COVID-19. However, at present, there is no consensus on the best diagnostic approach to predict outcomes in infected patients. Moreover, it is not clear whether such tools would apply to other potentially pandemic pathogens and therefore of use as stockpile for future pandemic preparedness.

**Methods:** We conducted a multi-cohort observational study to investigate the biology and the prognostic role of interferon alpha-inducible protein 27 (*IFI27*) in COVID-19 patients.

**Findings:** We show that *IFI27* is expressed in the respiratory tract of COVID-19 patients and elevated *IFI27* expression is associated with the presence of a high viral load. We further demonstrate that systemic host response, as measured by blood *IFI27* expression, is associated with COVID-19 severity. For clinical outcome prediction (e.g. respiratory failure), *IFI27* expression displays a high positive (0.83) and negative (0.95) predictive value, outperforming all other known predictors of COVID-19 severity. Furthermore, *IFI27* is upregulated in the blood of infected patients in response to other respiratory viruses. For example, in the pandemic H1N1/09 swine influenza virus infection, *IFI27-*like genes were highly upregulated in the blood samples of severely infected patients.

**Interpretation:** These data suggest that prognostic biomarkers targeting the family of *IFI27* genes could potentially supplement conventional diagnostic tools in future virus pandemics, independent of whether such pandemics are caused by a coronavirus, an influenza virus or another as yet-to-be discovered respiratory virus.

**Research in context:** *Evidence before this study:* We searched the scientific literature using PubMed to identify studies that used the *IFI27* biomarker to predict outcomes in COVID-19 patients. We used the search terms “*IFI27*”, “COVID-19, “gene expression” and “outcome prediction”. We did not identify any study that investigated the role of *IFI27* biomarker in outcome prediction. Although ten studies were identified using the general terms of “gene expression” and “COVID-19”, *IFI27* was only mentioned in passing as one of the identified genes. All these studies addressed the broader question of the host response to COVID-19; none focused solely on using *IFI27* to improve the risk stratification of infected patients in a pandemic.

*Added value of this study:* Here, we present the findings of a multi-cohort study of the *IFI27* biomarker in COVID-19 patients. Our findings show that the host response, as reflected by blood *IFI27* gene expression, accurately predicts COVID-19 disease progression (positive and negative predictive values; 0.83 and 0.95, respectively), outperforming age, comorbidity, C-reactive protein and all other known risk factors. The strong association of *IFI27* with disease severity occurs not only in SARS-CoV-2 infection, but also in other respiratory viruses with pandemic potential, such as the influenza virus. These findings suggest that host response biomarkers, such as *IFI27*, could help identify high-risk COVID-19 patients - those who are more likely to develop infection complications - and therefore may help improve patient triage in a pandemic.

*Implications of all the available evidence:* This is the first systemic study of the clinical role of *IFI27* in the current COVID-19 pandemic and its possible future application in other respiratory virus pandemics. The findings not only could help improve the current management of COVID-19 patients but may also improve future pandemic preparedness.

## INTRODUCTION

SARS-CoV-2 causes a broad range of clinical severity (mild to severe). What determines whether an infected individual will progress to severe COVID-19 is a complex interaction between host and viral factors. Since the beginning of the COVID-19 pandemic, there has been significant interest in developing robust biomarkers to predict disease outcomes amongst COVID-19 patients. This facilitates patient triage and resource prioritisation, both of which become particularly important when healthcare centres are required to simultaneously treat a large number of COVID-19 patients. Numerous prognostic indicators have been proposed for COVID-19. These include, but are not limited to, patient demographics, co-morbidities, lung computed tomography (CT) results, coagulation assays, alterations in white blood cell counts and inflammatory response biomarkers such as C-reactive protein and cytokines [1]. Others have used machine learning to predict disease outcomes [2, 3]. Despite these investigations, there is no consensus on the best approach for predicting COVID-19 outcomes. Moreover, as there is no indication as to whether such predictive tools would apply to other potentially lethal viruses, and therefore could become part of stockpile for future pandemic preparedness.

We have previously discovered that the transcription of the interferon alpha-inducible protein 27 (*IFI27*) is a signature marker of pandemic H1N1/09 influenza infection [4]. *IFI27* (also known as ISG12 or p27) is an interferon alpha (and to a lesser extent interferon gamma) inducible gene of unknown function, with the gene product residing in the nuclear membrane of the cell [5]. *IFI27* expression is associated with the severity of several different viral illnesses including respiratory syncytial virus (RSV) infection and Enterovirus 71 (EV71) hand foot and mouth disease [6, 7]. Preliminary evidence suggests that *IFI27* expression may be a useful biomarker for COVID-19 diagnosis and severity [8-11]. In the respiratory tract, *IFI27* expression is significantly upregulated by SARS-CoV-2 infection, more so than by other respiratory viruses [8, 9]. *IFI27* is also upregulated in the peripheral blood of COVID-19 patients [10, 11] and may serve as a biomarker for pre-symptomatic SARS-CoV-2 infection [12]. However, the usefulness of *IFI27* expression as a prognostic biomarker for COVID-19 remains to be determined.

Here, we use multiple patient cohorts to evaluate the role of *IFI27* expression in predicting COVID-19 disease progression. We further examine the role of *IFI27* expression for risk stratification in infections caused by other respiratory viruses, including influenza virus, another virus of pandemic potential.

## METHODS

Since *IFI27* is expressed in humans to varying degrees depending on the severity of COVID-19 infection, we assembled multiple patient cohorts (Cohort 1 - 10) to capture the full spectrum of clinical severity (‘Mild’, ‘Moderate’, ‘Severe’ as per definitions below). To ensure that population heterogeneity was adequately represented in the study, we recruited study participants across different countries (Brazil, Iran, Chile, Australia and the U.S.A.). To fully understand *IFI27* expression in different tissue compartments, we systematically evaluated samples of different tissues including lung, nasopharyngeal swabs, plasma and blood. A summary of the relevant cohort characteristics and sampling methods is provided in Table 1. The study was approved by the Human Research Ethics Committees of participating institutions and all study participants provided informed consent.

**Table 1:** Overview of cohorts included in the study.

|  | Country | Year | Study design | Pathogen identified | Patients recruited | Healthy controls | Gender (males/females) | Age (Mean years) | Tissue type | Experiment performed | *IFI27* measurement | Clinical setting | Disease severity® | Admission to hospital (%) | Death (%) |
| --- | --- | --- | --- | --- | --- | --- | --- | --- | --- | --- | --- | --- | --- | --- | --- |
| Cohort 1 | Brazil | 2020 | Case-control study | SARS-CoV-2 | 10 | 4 | 8/2 | 64 | Lung | RNAscope & Spatial transcriptomics | 10X Genomics Visium | Hospital | Severe | 10 (100%) | 10 (100%) |
| Cohort 2 | Chile | 2020 | Cross-sectional study | SARS-CoV-2 | 137 | 0 | 92/45 | 46 | Nasal Swabs | qPCR | qPCR | Hospital | Moderate Severe | 130 (100%) | NA |
| Cohort 3 | U.S.A. | 2020 | Cross-sectional study | SARS-CoV-2 | 60 | 0 | 30/30 | 55 | Nasal Swabs | qPCR | qPCR | Outpatient and hospital | Mild Moderate Severe | 38 (63%) | 4 (6.7%) |
| Cohort 4 | Chile | 2020 | Cross-sectional study | SARS-CoV-2 | 127 | 0 | 83/44 | 54 | Plasma | ELISA | ELISA | Hospital | Moderate Severe | 127 (100%) | 16 (12.5%) |
| Cohort 5 | Brazil | 2020 | Cross-sectional study | SARS-CoV-2 | 28 | 6 | 18/10 | 60 | Plasma | ELISA | ELISA | Hospital | Mild Moderate Severe | 19 (68%) | 4 (14%) |
| Cohort 6 | Iran | 2020 | Cross-sectional study | SARS-CoV-2 | 16 | 6 | 7/9 | 53 | Blood | qPCR | qPCR | Hospital | Asymptomatic, Mild Moderate | 12 (75%) | 4 (25%) |
| Cohort 7 | Australia & Singapore | 2020 | Prospective cohort | SARS-CoV-2 | 44 | 14 | 18/26 | 49 | Blood | qPCR | qPCR | Community, outpatient, and hospital | Mild Moderate Severe | 24 (55%) | 0 (0%) |
| Cohort 8 | Australia | 2021 | Cross-sectional study | SARS-CoV-2 (Delta) | 22 | 13 | 13/9 | 40 | Blood | qPCR | qPCR | Hospital | Moderate Severe | 22 (100%) | 0 (0%) |
| Cohort 9 | Australia | 2013 - 2018 | Prospective cohort | Influenza, RSV, others# | 228 | 13 | 120/108 | 51 | Blood | qPCR | qPCR | Community, outpatient and hospital | Mild Moderate Severe | 137 (60%) | 8 (4%) |
| Cohort 10 | Australia | 2009 | Cross-sectional study | Influenza (H1N1/09 swine) | 107 | 52 | 44/63 | 49 | Blood | Microarray | Microarray | Community, outpatient, and hospital | Mild Moderate Severe | 89 (83%) | 9 (8.4%) |
| Total |  |  |  |  | 779 | 108 |  |  |  |  |  |  |  |  |  |

@ For disease severity, ‘Mild’ disease is defined as the presence of COVID-19 disease (as confirmed by the admitted clinician) in a patient who does not require hospitalization. ‘Moderate’ disease is defined as the presence of COVID-19 disease in a patient who requires hospitalization but does not need mechanical ventilation. ‘Severe’ disease is defined as the presence of COVID-19 disease in a patient who requires both hospitalizations and mechanical ventilation (usually in intensive care unit). # ‘Others’ denotes other respiratory viruses such as parainfluenza virus, rhinovirus, humanmetapneumovirus. qPCR denotes ‘quantitative polymerase chain reaction’. ELISA denotes ‘enzyme-linked immunosorbent assay’.

### Definition of severity

We adopted a simplified version of the CDC definition of COVID-19 disease severity (13). In this simplified definition, COVID disease was defined as the presence of suspected COVID-19 symptoms (e.g., fever, sore throat) (Supplementary Table 1), together with a positive SARS-CoV-2 detection using virus nucleic acid amplification assay (qPCR). ‘Mild’ disease was defined as the presence of COVID-19 disease (as confirmed by the admitting clinician) in a patient who did not require hospitalization. ‘Moderate’ disease was defined as the presence of COVID-19 disease in a patient who required hospitalization. ‘Severe’ disease was defined as the presence of COVID-19 disease in a patient who required mechanical ventilation (in an intensive care unit).

### Study design

We deployed both retrospective and prospective studies to understand the biology of *IFI27* and to evaluate the clinical utility of the *IFI27* biomarker. Cohort 1 was a case-control study in which the biological role of *IFI27* expression in SARS-CoV-2-induced acute lung injury was examined. Cohorts 2-6 were cross-sectional studies. We used these studies to compare *IFI27* expression across different tissue compartments (serum, airways and blood), in order to determine the most suitable sampling route for *IFI27* measurement. Cohort 7 and 9 were prospective cohorts used to validate the prognostic performance of the *IFI27* gene-expression biomarker in a real-world setting. Cohorts 9 and 10 extended the analysis to other respiratory tract virus infections, including the influenza virus which is another pathogen with pandemic potential. Table 1 provides a summary of all cohorts included in this study. Additional details on patient recruitment, experiments performed, and data collection are also provided in the **Supplementary Methods**.

### Outcome measure

Cohort 7 was a prospective validation study of *IFI27* in predicting COVID-19 outcomes. As the sample size was small (n=44), a composite outcome was used to evaluate *IFI27* prediction performance. The composite outcome in a COVID-19 patient was defined as, during the 28-day study period, the first occurrence of: (1) any complication as defined by the International Severe Acute Respiratory and Emerging Infection Consortium (ISARIC), such as viral pneumonia, acute respiratory distress syndrome (ARDS) or bacterial pneumonia (Supplementary Table 1); or (2) prolonged virus shedding; or (3) ICU admission; or (4) hospital stay > 7 days. A patient with a ‘poor’ outcome was one in whom a composite outcome occurred within the 28-day study period and a patient with a ‘good’ outcome was one in whom a composite outcome had not occurred during the 28-day study period.

### Predictive performance

We assessed *IFI27* predictive performance using the established methods of Metz and Zhou, as implemented in the NCSS statistical software (Utah, U.S.A) [14]. Sensitivity, specificity, positive predictive values, negative predictive values, positive likelihood ratio and negative likelihood ratio were calculated using the previously established cut-off value for *IFI27* (74) [4]. For all performance metrics, 95% confidence intervals were calculated based on the Exact (Clopper-Pearson) method. [15]. For assessing predictive performance, a previously validated cut-off threshold for *IFI27* expression (fold-change of 74) was used [4]. For grouping ‘mild’, ‘moderate’ and ‘severe’ COVID-19 patients, clinical criteria (see ‘Definition of severity’) were used for this purpose.

### Experiments

(1) *<u>cDNA synthesis and qPCR</u>*; Total RNA was reverse transcribed using a Qscript cDNA SuperMix (QuantaBio). Amplification of *IFI27* was performed using TaqMan gene expression Master Mix on a CFX384 system. GAPDH was used as the endogenous control. The delta-CT method was used to calculate the fold change in gene expression. (2) *<u>ELISA</u>*; Plasma samples, collected from COVID-19 patients were analysed in duplicate using the IFI-27 ELISA kit (Aviva Systems Biology, USA). Plasma IFI-27 (pg/ml) was log2 transformed and median centred to evaluate the relationships to clinical outcomes of COVID-19. (3) *<u>RNAscope</u>*; RNAscope^®^ probes (Advanced Cell Diagnostics, USA) targeting SARS-CoV-2 spike mRNA were used according to manufacturer’s instructions for automation on the Leica Bond RX of formalin-fixed paraffin embedded (FFPE) rapid autopsy lung tissues from COVID-19 patients and controls. Fluorescent images were acquired with Nanostring Mars prototype DSP at 20x. (4) *<u>Spatial transcriptomics</u>*; FFPE samples were sectioned at 7µm thickness using a microtome and the section was transferred to a water bath at 41°C. The floating section was adhered to the Visium Spatial Gene Expression Slide (10x Genomics, USA) and processed as per manufacturer recommendations. (5) *<u>Microarray</u>*: The microarray data of GSE101702, which included 107 influenza patients (n=63 moderate and n=44 severe) and 52 healthy controls, was analysed. The clinical characteristics and additional detail of the dataset are previously described [16]. We identified the differentially expressed genes (DEGs) by the R package ‘limma’ [17] between moderate influenza and severe influenza samples. Genes with 1 log2fold change with adjusted P value < 0.05 (0.05 FDR) value were considered significant. Full details on the above methods are also provided in **Supplementary Methods**.

### Statistical analysis

Data were tested for normality using the Anderson-Darling test. Where data were normally distributed, they were analysed using an unpaired two-tailed student’s t-test or a one-way ANOVA with a Holm-Šídák’s multiple comparisons test. Where data were not normally distributed, they were analysed using the Mann-Whitney U test or a Kruskal-Wallis test with Dunn’s multiple comparison test. The significance was set at p<0.05. All statistical analyses were performed using Prism version 9.0.

## RESULTS

The findings of this study included expression data (gene/protein) generated from 779 patients and 108 healthy controls, assembled in ten cohorts across six countries (Australia, U.S.A., Chile, Brazil, Iran, and Singapore). There was one case-control study (Cohort 1), two prospective studies (Cohorts 7 and 9) and seven cross-sectional studies (Cohorts 2, 3-6, 8 and 10). Patients included in the studies were drawn from different clinical settings (community, outpatient clinics and hospitals) and disease severities (mild, moderate and severe). Cohort 8 included COVID-19 patients recruited in 2021, who were infected exclusively by the delta-variant virus of SARS-CoV-2 (as confirmed by full genome sequencing). Other COVID-19 cohorts recruited patients in early 2020 (Cohorts 1-7). In these early cohorts, no genome sequence data were available (due to limited access to genome sequencing facilities) to identify virus variant subtypes. Several methods were used to measure *IFI27* gene expression including PCR, microarray, and spatial transcriptomics. Table 1 provides full details of the *IFI27* measurement methods and tissue sampling approaches in each cohort.

We first investigated the biological role of *IFI27* in COVID-19 by assessing gene expression in the lower respiratory tract of deceased COVID-19 patients (Cohort 1; n=10). The virus load in the lung was assessed by RNAscope^®^, which had the sensitivity to detect single molecules in a cell. The distribution of virus load was quantified by STRISH, a robust image processing pipeline. The average measurements of SARS-CoV-2 spike mRNA (nCoV2019) per grid (tissue region) of neighbouring cells were visualized by STRISH using a tissue heatmap (Figure 1A). These data were processed by the Visium^®^ spatial transcriptomic method to unbiasedly profile ∼22,000 genes across thousands of spatial distributed spots within tissue sections. Based on these analyses, we discovered that areas of high viral load in the lung had high levels of *IFI27* gene expression (Figure 1B, C). The correlation between high viral load and increased *IFI27* gene expression was independently replicated by two different Visium^®^ spatial protocols, one with polyA-capture (method 1) and another with probe hybridization (method 2). We also applied the same analysis to the lung samples of age and sex-matched control patients (n = 8). As expected, neither *IFI27* nor other immune response genes were expressed in healthy lung tissue or when the SARS-CoV-2 virus was absent (data not shown).

**Figure 1:**
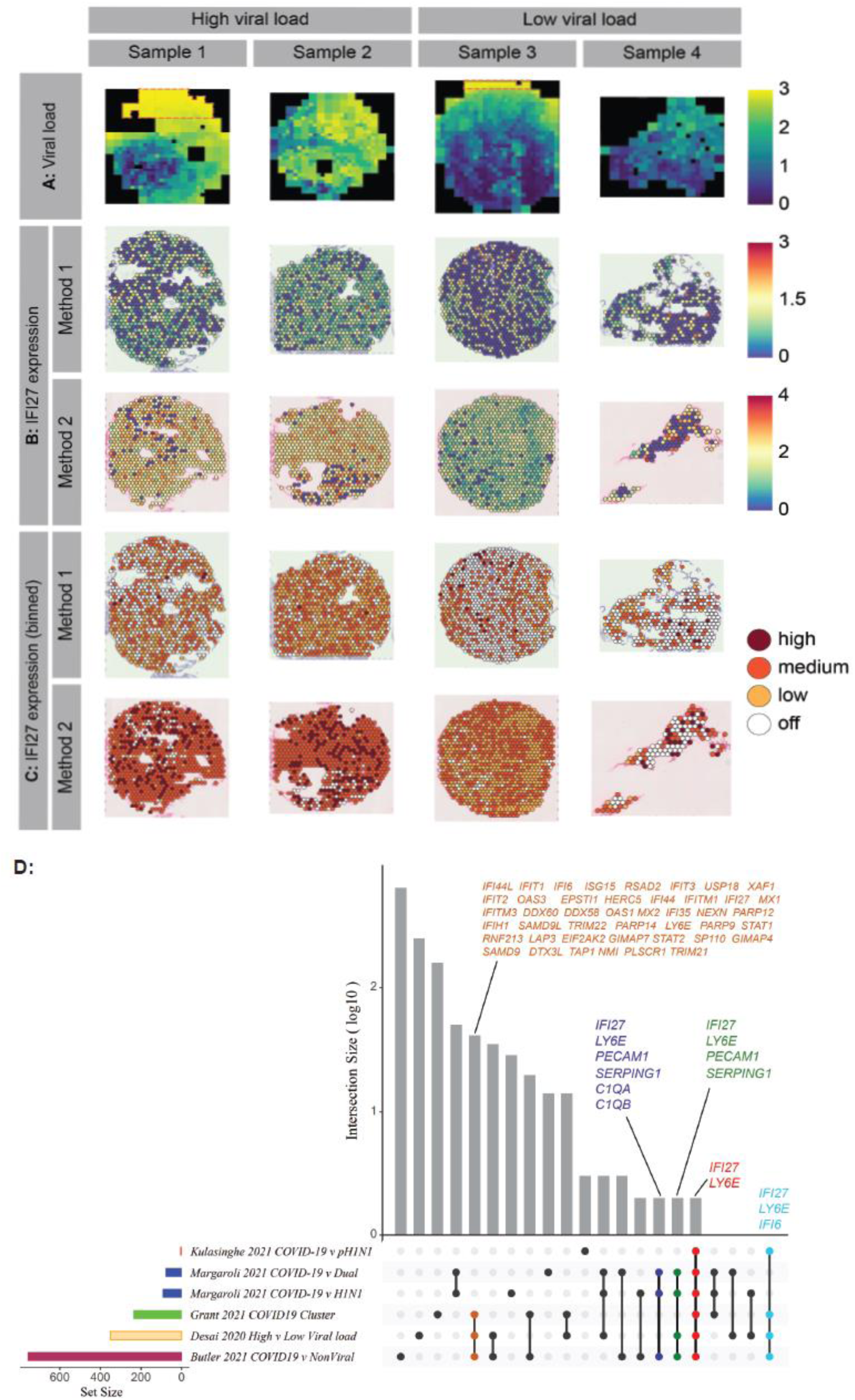
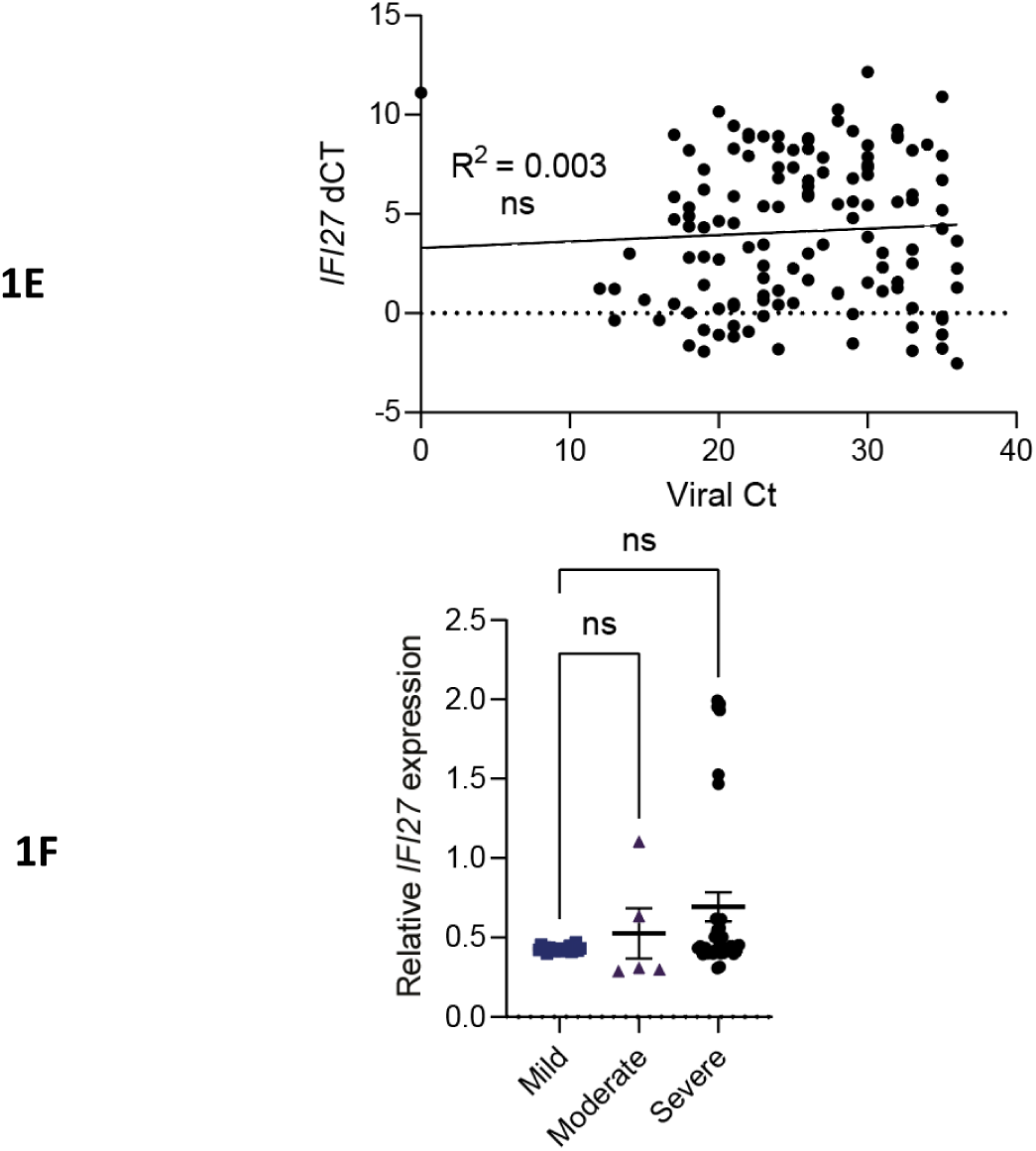
Airway *IFI27* gene expression in COVID-19 patients. **A**) In Cohort 1, spatial expression heatmap of normalised COVID-19 RNAscope signal was determined using STRISH analysis. The heatmap colour shows average RNAscope signal per cell per rectangle area that contains a similar number of cells (fewer than 100 cells per rectangle). Red boxes indicate areas of overexposure in the original RNAscope microscopy, which were excluded from the analysis. **B)** Normalised expression of *IFI27* across four samples measured in the same Visium experimental slide was shown (blue as low and yellow-red as high). Results from two different methods run separately by two labs and applied for the same tissue block are shown. Method 1 and Method 2 correspond to the poly-A protocol and the 10x capture protocol as described in the Methods. **C)** Binned values of normalised expression of *IFI27* across Visium samples. **D)** Upset plot describing the overlapping gene sets across 5 studies. The size of the gene set varies across the studies with a small number of common genes including *IFI27* shared across all studies (18-20). **E)** *IFI*27 gene expression in nasopharyngeal samples in Cohort 2 (n=137). SARS-CoV-2 virus load (as measured by Ct values) is used as a proxy of local disease activity. A statistically non-significant *p*-value of the linear regression model (represented by R^2^) indicates that there is no association between *IFI*27 expression and virus load or local disease activity. **F**) *IFI*27 gene expression in nasopharyngeal samples in Cohort 3 (n=60). ‘Mild’ disease is defined as the presence of COVID-19 disease in a patient who does not require hospitalization. ‘Moderate’ disease is defined as the presence of COVID-19 disease in a patient who requires hospitalization. ‘Severe’ disease is defined as the presence of COVID-19 disease in a patient who requires mechanical ventilation in an intensive care unit. p value is calculated using Mann-Whitney U test. * p <0.05; ns = not significant. Data shows mean ± SEM. *IFI*27 gene expression is measured by qPCR normalized to house-keeping genes.

Further analysis of the expression of an additional 488 host genes (from the same region of the lung samples) showed that *IFI27* was co-expressed with other host genes involved in anti-viral response (Supplementary Table 2). This indicated that *IFI27* expression was part of an intra-pulmonary immune response against SARS-CoV-2. This finding was validated by several recently published studies (18-20), which not only confirmed the upregulation of *IFI27* expression in infected lung tissue of COVID-19 patients, but also showed that *IFI27* was the most consistently expressed host response gene across all studies (Figure 1D).

**Table 2.** Performance of blood *IFI27* gene-expression in COVID-19 outcome prediction

| Parameters | Sensitivity | Specificity | Positive predictive value | Negative predictive value | AUROC |
| --- | --- | --- | --- | --- | --- |
| *IFI27* | 0.95 | 0.83 | 0.83 | 0.95 | 0.90* |
| Lymphopenia | 0.40 | 0.71 | 0.48 | 0.64 | 0.60 |
| CRP | 0.72 | 0.75 | 0.66 | 0.80 | 0.69 |
| Age | 0.50 | 0.54 | 0.42 | 0.62 | 0.57 |
| Comorbidity | 0.70 | 0.64 | 0.57 | 0.76 | 0.77* |
| Symptom score | 0.80 | 0.50 | 0.52 | 0.79 | 0.74* |
| Heart rate<sup>¶</sup> | 0.65 | 0.73 | 0.61 | 0.76 | 0.71* |
| Respiratory rate<sup>¶</sup> | 0.45 | 0.91 | 0.77 | 0.71 | 0.59 |
| PaO<sub>2</sub><sup>¶</sup> | 0.42 | 0.64 | 0.43 | 0.62 | 0.53 |

AUROC denote area under the curve of receiver-operating-characteristic curve. \* Statistically significant (p<0.01).
<sup>¶</sup> These measurements (heart rate, respiratory rate and PaO<sub>2</sub>) were performed on the sample time where blood samples were taken for *IFI27* measurement. This is usually done immediately after the patient was recruited into the study (within 24 hours of each patient's initial presentation). If there were multiple measurements within the same 24 hours period, by convention, the worst reading was recorded.

The above data indicated that *IFI27* gene expression may reflect local disease activity in infected lung tissue. However, lower respiratory tract is not readily accessible or available for routine diagnostics. Accordingly, the association between *IFI27* gene expression and disease severity was assessed using upper airway samples (nasopharyngeal swabs) of COVID-19 patients (Cohort 2). Here, no significant association was observed between nasopharyngeal *IFI27* gene expression and virus loads (Figure 1E). Further analysis in another cohort (Cohort 3) showed a similar finding; there was no difference in *IFI27* gene expression between mild, moderate and severe disease (Figure 1F). Since these findings suggested that upper airway *IFI27* expression levels did not reflect disease activity in the lower airway, we therefore sought an alternative sampling route to measure *IFI27* expression in COVID-19 patients.

The spread of SARS-CoV-2 infection to the lower airway is often associated with a systemic host response, and blood provides an easily accessible clinical sample to assess this response. Thus, we proceeded to measure *IFI27* levels in peripheral blood of COVID-19 patients (Cohort 6). We found that blood *IFI27* was higher in infected patients (compared to asymptomatic or uninfected individuals). Furthermore, there was a trend of increasing *IFI27* gene expression in patients with a worsening disease, although this trend was not statistically significant (Figure 2A). Similarly, protein expression of *IFI27* was also elevated in COVID-19 patients (Cohorts 4 and 5). However, *IFI27* protein expression did not correlate with disease severity (Figure 2B). These findings suggested that blood *IFI27* gene expression (not protein expression) could act as a surrogate marker of disease severity in COVID-19.

**Figure 2:**
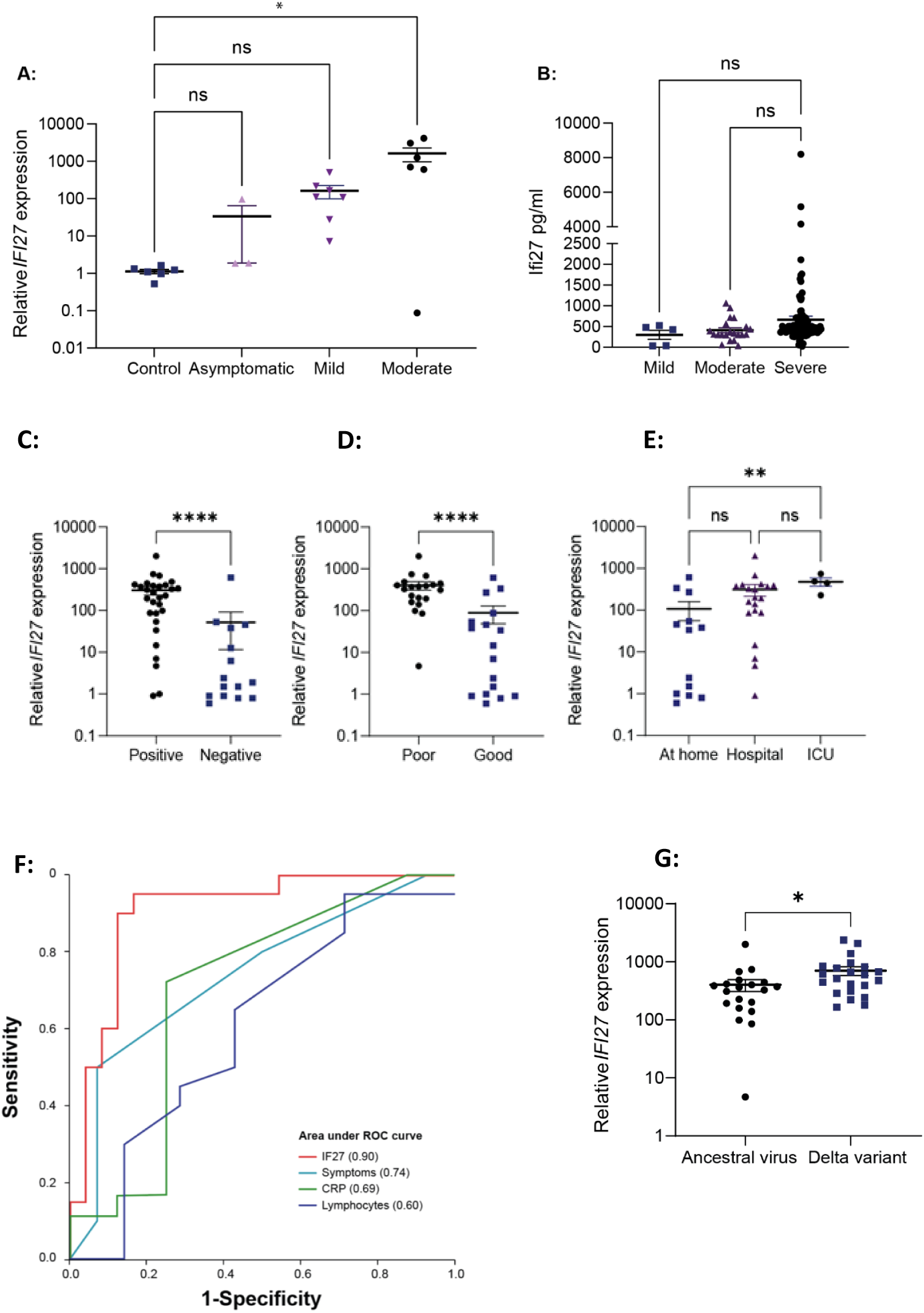
Blood *IFI27* gene expression in COVID-19 patients. **A)** Blood *IFI27* gene-expression levels in healthy controls, uninfected and infected patients in Cohort 6 (n =16). **B)** Blood *IFI27* protein-expression levels in Cohorts 4 and 5 (n=161). *IFI*27 protein-expression and statistical significance was determined as described in the Methods. **C)** Blood *IFI27* expression in Cohort 8, between those actively shedding viruses (labelled as ‘positive’) and those not shedding virus patients (labelled as ‘negative’). **D)** Blood *IFI27* gene expression in Cohort 7 (n=44) for prospective validation of outcome prediction. Patients were grouped under ‘poor’ if an adverse outcome has occurred, or ‘good’ if an adverse outcome has not occurred. **E)** Blood *IFI27* gene expression in Cohort 7 (n=44) in patients with different dispositions. ICU denote ‘intensive care unit’. p value is calculated using Mann-Whitney U test. ** p <0.01; ****p<0.0001; ns = not significant. Data shows mean ± SEM. **F:** Area-under-the-curve of Receiver-Operator-Characteristics curve (AUROC) analysis of blood *IFI27* gene expression levels as a predictor of clinical outcome in Cohort 7 (n=44). *IFI27* gene-expression level (‘IFI27’), total number of symptoms (‘Symptoms’), C-reactive protein (‘CRP’) and lymphocyte count (‘Lymphocytes’). **G:** Blood *IFI27* in SARS-CoV-2 Delta-variant (Cohort 8, n=22) and ancestral (non-Delta variant) (Cohort 7, n=44). p value is calculated using Mann-Whitney U test. *p <0.05; ns = not significant. Data shows mean ± SEM.

We used an independent prospective cohort (Cohort 7) to test the hypothesis that changes in blood *IFI27* gene expression could predict outcomes in COVID-19 patients. In Cohort 7, blood sampling (for *IFI27* gene expression) was performed upon initial presentation of each patient when the disease outcome was still unknown. Each patient was then followed up for 28 days and their clinical outcomes (e.g., acute respiratory distress syndrome) were recorded (see Methods). We found that blood *IFI27* gene expression was significantly higher in patients with progressive disease, such as those patients in whom active virus shedding was ongoing (Figure 2C), who developed an adverse outcome (Figure 2D) or were admitted to the intensive care unit (Figure 2E). In patients who were admitted to the intensive care unit, the *IFI27* gene expression increase could precede - by several days - clinical signs of deteriorations or abnormal changes in laboratory parameters such as high levels of C-reactive protein (Supplementary Figure). This observation was further confirmed by area under the curve of receiver-operator-characteristics curve (AUROC) analysis, which showed the *IFI27* gene expression outperformed laboratory variables (e.g., C-reactive proteins), patient variables (e.g., age, comorbidity) and physiological parameters (e.g., respiratory rate) in predicting COVID-19 outcomes (Table 2 and Figure 2F). Notably, the *IFI27* gene expression had a high sensitivity (0.95), high specificity (0.83) and an AUROC of 0.90, all of which were higher than known factors associated with COVID-19 outcomes (e.g., age, co-morbidity). Given that patients Cohort 7 was recruited in early 2020 (Table 1), patients from a recent delta-variant outbreak (Cohort 8) were also recruited. In Cohort 8, we found a similar association between an increased *IFI27* gene expression levels and severe disease (Figure 2G).

In Cohort 9 (n=228), we compared blood *IFI27* gene expression across different respiratory infections, including both bacterial and viral infections (Figure 3A) (details of Cohort 9 had been previously described [4]). This expanded analysis showed that bacterial respiratory infection did not result in elavated *IFI27* gene expression. In contrast, most respiratory viruses upregulated *IFI27* expression (Figure 3A). Notably, *IFI27* upregulation is the highest in influenza and SARS-CoV-2 infection. Given the importance of the influenza virus as a pandemic virus, we further analysed blood *IFI27* gene expression data from a previously published microarray study [16]. This analysis confirmed that *IFI27* upregulation occurred in severe influenza infection (Figure 3B). Importantly, the *IFI27* gene family-including interferon alpha-inducible protein 27-like protein 1 (*IFI27L1*) and interferon alpha-inducible protein 27-like protein 2 (*IFI27L2*)-were better at discriminating disease severity (Figure 3C, 3D). Together, these expanded analyses demonstrated that *IFI27*, or its gene family, could be a signature marker of the host response to respiratory viruses, and that an increasing *IFI27* upregulation could be a warning sign of infections by more lethal viruses.

**Figure 3:**
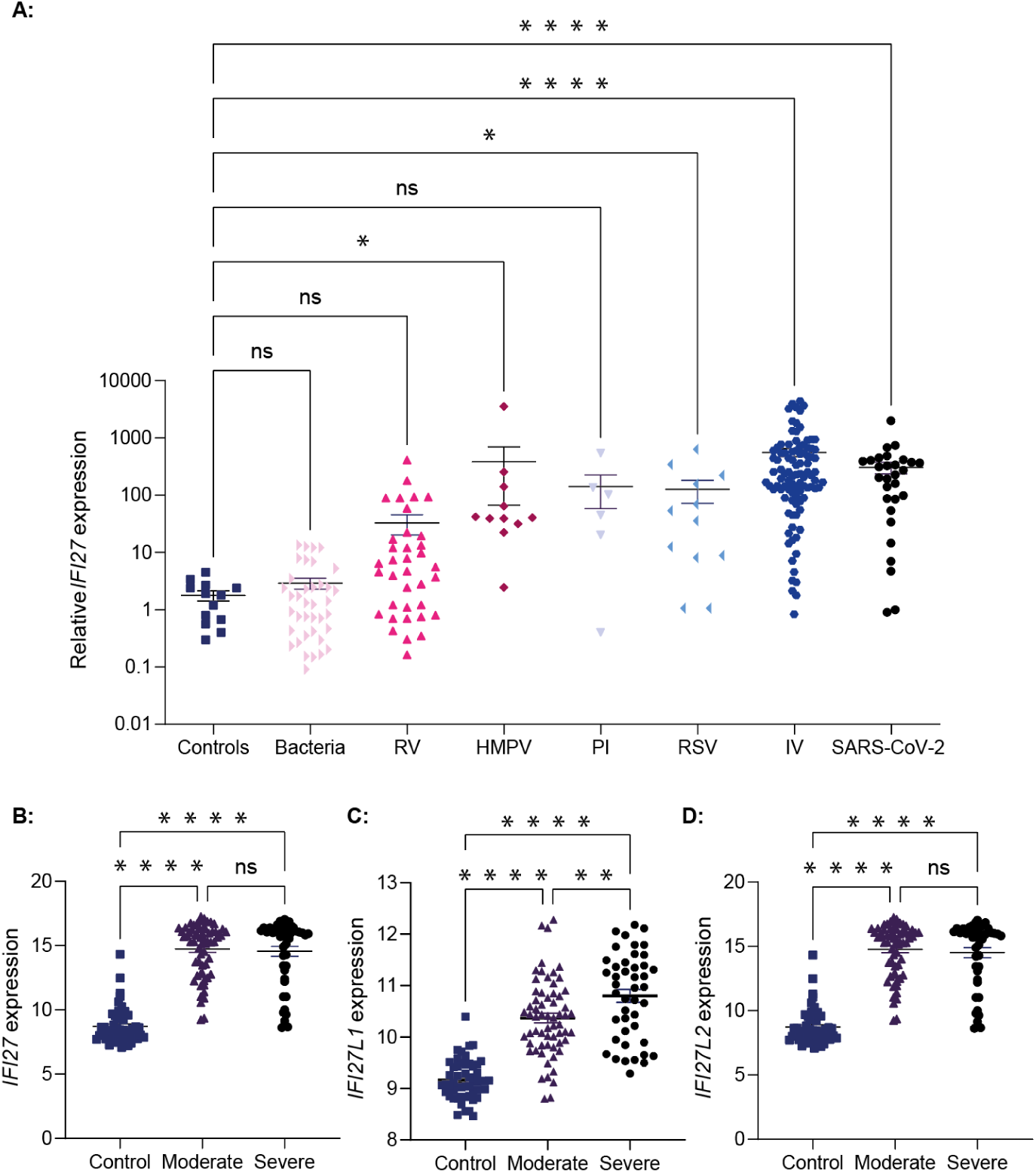
Expression of *IFI27* gene family members in different respiratory infections. **A)** Blood *IFI27* gene expression in different infectious disease aetiologies in Cohort 9 [4], which includes infected baseline controls (‘control’ n = 13), bacterial respiratory infection (‘bacteria’ n = 38), rhinovirus (‘RV’ n = 36), human metapneumovirus (‘HMPV’ n = 11), parainfluenza (‘PI’ n = 6), respiratory syncytial virus (‘RSV’ n = 12), influenza A or B virus (‘IV’ n = 96) or SARS-CoV-2 (n = 29). *IFI*27 gene expression was measured by qPCR normalized to house-keeping genes. **B-D)** Gene expression in the blood of 107 influenza patients. ‘Mild’ disease is defined as the presence of COVID-19 disease in a patient who does not require hospitalization. ‘Moderate’ disease is defined as the presence of COVID-19 disease in a patient who requires hospitalization. ‘Severe’ disease is defined as the presence of COVID-19 disease in a patient who requires mechanical ventilation in an intensive care unit. p value is calculated using Mann-Whitney U test. * p <0.05; ** p <0.01; **** p<0.0001; ns = not significant. Data shows mean ± SEM.

## DISCUSSION

Globally, more than 400,000 new SARS-CoV-2 infections are recorded every day and the emergence of novel viral variants has raised concerns that these numbers will continue to increase, despite the increasing availability of vaccines. The current global situation emphasises the ongoing need for COVID-19 prognostic biomarkers to facilitate both patient triage and resource prioritisation. Here, we have provided the first evidence of blood *IFI27* expression as a potential biomarker for risk stratification of COVID-19 patients. When prospectively validated, blood *IFI27* expression showed a high positive and negative predictive value, outperforming other known predictors of COVID-19 severity reported in the literature.

*IFI27* is an interferon inducible gene. Accordingly, the strong association observed between *IFI27* upregulation and severe COVID-19 could be indicative of an increased viral replication. However, this hypothesis would be inconsistent with the poor correlation observed between nasopharyngeal *IFI27* expression and virus load. Instead, we propose that *IFI27* expression reflects increased immunopathology (either local or systemic) and is thereby associated with COVID-19 severity. The correlation of interferon expression and the severity of COVID-19 shows significant anatomical variation. In the upper respiratory tract, rapid induction of type I interferons is typically associated with reduced COVID-19 severity, as their induction is associated with the ability to control viral replication with limited immunopathology (21,22). In contrast, in the lower respiratory tract, the induction of type I interferons is associated with an exacerbated inflammatory response and significant tissue damage (23). Similarly, the systemic induction of type I interferons is associated with immunopathology and distal tissue damage (24). Therefore, we hypothesise that blood *IFI*27 expression was associated with severe disease and that blood *IFI27* expression had prognostic value (in contrast to *IFI27* levels in upper airway samples). Importantly, we observed a stronger association between COVID-19 outcome and the *IFI*27 gene expression than that observed with the *IFI27* protein expression in blood. This most likely reflects an increased dynamic range for measuring gene expression and thereby an increased ability to differentiate patient outcomes. Future attempts to translate these findings into routine clinical settings should therefore focus on PCR-based assays to measure *IFI27* expression in blood.

Compared to other biomarkers reported in COVID-19 literature, *IFI27* offers several clinical advantages. Firstly, *IFI27* expression appears to be specific to viral illness. In contrast, most infection/inflammatory biomarkers (e.g., C-reactive protein, leukocytes, interlukin-6) are elevated in many non-viral illnesses (e.g., trauma, sepsis). Secondly, *IFI27* expression is directly linked to intra-cellular recognition of respiratory viruses (21-24). Given the control point of *IFI27* expression lies on the disease causal pathway, it makes sense to track disease progression by using *IFI27* expression, rather than via biomarkers that are unrelated to the underlying disease activity. Finally, blood *IFI27* gene expression has a strikingly high dynamic range (e.g., up to thousands of fold changes in severe COVID-19, as shown in the present study) and a strong signal-to-noise ratio (which produced consistent findings across different measurement platforms, such as microarray or PCR, also evidenced in our findings). These favourable measurement characteristics makes *IFI27* a preferred prognostic tool to other biomarkers.

The kinetics of *IFI27* expression is poorly understood. As a result, the ideal sampling window was not well-defined in this study. It is also uncertain whether serial measurement of *IFI27* expression would be more informative than a single time point measurement. Furthermore, the predictive performance of *IFI27* could be confounded by other yet-to-be-defined variables, such as timing (e.g., early presentation versus late presentation), stages of disease (e.g., lung only infection versus multi-organ disease) and COVID-19 prevalence. For these reasons, we are unable to extrapolate the prognostic value of blood *IFI27* expression to a broader clinical context; additional studies are required. Such studies should consist of large-scale, prospective studies with sample sizes adequately powered to allow researchers to assess the independent confounding effect of each variable (e.g., timing, disease stages and prevalence) on *IFI27* expression.

The COVID-19 pandemic has emphasised the need to have stockpiles of broad-spectrum diagnostic and therapeutic tools that can be rapidly deployed at the start of any future viral outbreak. As a component of the anti-viral interferon response, it is not surprising that elevated *IFI27* expression was observed in the blood of patients infected with a broad range of different respiratory viruses but not in the blood of patients with a bacterial respiratory infection. These data, combined with previous suggestions that *IFI27* expression is associated with the severity of RSV [6, 7], suggest that *IFI27* may represent a pan-viral prognostic biomarker. Interestingly, when we investigated this further in the context of influenza virus, we found that expression of *IFI27L1* in the blood, rather than *IFI27* itself, was associated with disease severity. We therefore propose that future studies focus on developing PCR-based assays to measure a suite of genes associated with *IFI27* expression including *IFI27L1, IFI27L2* and *IFI6*. It is hoped that this broader, combinatorial approach, would lead to the development of pan-viral prognostic biomarker that could be incorporated into future pandemic preparedness planning.

The present study has several limitations. Firstly, the prospective validation cohort was relatively limited in size. Secondly, patient cohorts were recruited prior to the availability of vaccination. Accordingly, our findings do not apply to vaccinated individuals experiencing ‘breakthrough’ infections. Finally, we could not correlate the changes in *IFI27* expression levels between the peripheral blood and the infected lung. Future studies (e.g., in animal models) are needed to unravel the coupled dynamics of *IFI27* expression between these tissue compartments.

In conclusion, the findings provided herein represent the first evidence that *IFI27* expression has a potential as a severity biomarker for risk stratification in COVID-19 patients.

## Data Availability

All data produced in the present study are available upon reasonable request to the authors

## Acknowledgements

We would like to acknowledge the following institutions/individuals: L. Pan, A. Nam (Nanostring Technologies, Seattle, USA), T. Y. Drennon, C. R. Uytingco, S. R Williams (10X Genomics, Pleasanton, USA), Nepean Institute of Critical Care Education and Research and Westmead Scientific Platforms supported by Westmead Institute for Medical Research, Cancer Institute New South Wales and the National Health and Medical Research Council. Tania Sorrell and Sue Maddock for their support of the study in Cohort 7 and patients and families that made this study possible.

## Author Contributions

AK, KS, BT, AM, MS conceived the study. AS, JM, LC, MT, GRR, TRM, MN, KYC, YZ, YX, TJW, ACS, CLKR, CLF, AFRDS, LDN, SM, RV, OA, EM, RAN, LL, FZ, ENL, GL, BK, VH, AP, TC, TK, KK, ST, TMP, WSK, YJ, JI performed the experimentation. CWT, RG, YW, KOB, JFF, MJD, GB, MEW, CSG, FSGG, QN performed the data analysis and interpretation. All authors critically reviewed and approved the manuscript for submission

## Supplementary File

Supplementary Methods

Supplementary Figure

Supplementary Table 1

Supplementary Table 2

## SUPPLMENTARY METHODS

### Cohort 1

Tissue microarray cores were prepared from autopsied pulmonary tissue from SARS-CoV-2 patients who died from respiratory failure (ARDS). Control material was obtained from 4 uninfected patients. All SARS-CoV-2 patients were confirmed for infection through RTqPCR of nasopharyngeal swab specimens, and imaging with computed tomography (CT) showed diffuse and bilateral opacities with ground-glass attenuation, suggestive of viral pulmonary infection. Autopsy and biopsy materials were obtained from the Pontificia Universidade Catolica do Parana PUCPR the National Commission for Research Ethics (CONEP) under ethics committee approval reference number 2020001792/30188020.7.1001.0020 and approval reference number 2020001934/30822820.8.000.0020. The study was also approved under University of Queensland Human Research Ethics Committee (HREC) ratification.

### Cohort 2 & 4

This study was approved by the Human Research Ethics Committees of the University of Concepcion (Chile) (CEBB 676-2020) and ratified by the University of Queensland (2021/HE000319). All methods were performed in accordance with institutional guidelines and regulations. Written consent was obtained from all study participants. Study participants were recruited from Hospital Regional Dr. Guillermo Grant Benavente, Concepcion, Chile. The inclusion criteria were nasopharyngeal swab RT-PCR confirmed SARS-CoV-2 infection. Nasal samples (n=137) and blood samples (n=127) were collected. Nasal samples (n=137) were grouped as “Cohort 2” and blood samples (n=127) were grouped as “Cohort 4”. COVID-19 disease severity at presentation to hospital was recorded for the patient group from which the blood samples were obtained. Mild (n=3), Moderate (n=16) and severe (n=108). “Mild” disease is defined as the presence of COVID-19 disease in a patient who does not require hospitalization. “Moderate” disease is defined as the presence of COVID-19 disease in a patient who requires hospitalization. “Severe” disease is defined as the presence of COVID-19 disease in a patient who requires mechanical ventilation in an intensive care unit.

### Cohort 3

The COVID-19 samples in form of the nasopharyngeal swabs were procured from the TSB BioBank, which is part of the Translational Science BioCore (TSB) affiliated with the UW Carbone Cancer Center (UWCCC), University of Wisconsin-Madison School of Medicine and Public Health, Madison, Wisconsin, USA. Coronavirus Disease 2019 (COVID-19), PCR (UWH) test was performed on nasopharyngeal swabs on Molecular Genprobe Panther Fusion platform in Molecular Diagnostics Lab. Chart review was performed on admitted patients negative for COVID19 and patients who presented to ED with symptoms consistent with COVID19 infection, such as fever, cough, and dyspnea were included in the cohort. Chart review was performed on admitted patients positive for COVID19 and patients who had only 1 positive COVID test result in their clinical history on the day of their admission were included in the cohort.

### Cohort 5

This study has been approved by Human Research Ethics Committee of the Pontifícia Universidade Católica do Paraná (PUCPR) (Number CAAE: 30833820.8.0000.0020) and by Comissão Nacional de Ética em Pesquisa (CONEP). All methods were performed in accordance with institutional guidelines and regulations. Written consent was obtained from all study participants. Study participants were recruited from Complexo Hospital de Clinicas da Universidade Federal do Paraná who had tested positive by PCR for SARS-CoV-2. Subjects with previous or current history of malignancy; thromboembolic pathology; severe allergic reaction; concomitant infection with HIV, tuberculosis, or other respiratory virus; transplant or use of immunosuppressive therapy and pregnancy or breastfeeding were not included in the study. Patient information is listed in Table 1.

### Cohort 6

This study was approved by the Mazandaran University of Medical Sciences (approval number IR.MAZUMS.REC 1399.856). All methods were performed in accordance with institutional guidelines and regulations. Written consent was obtained from all study participants. The inclusion criteria were nasopharyngeal swab RT-PCR confirmed SARS-CoV-2 infection and computed tomography (CT) scan with ground-glass opacities. A subgroup of participants without SARS-CoV-2 infection were included as controls. Patients (n=16) and uninfected individuals (n=6) were recruited for this study and provided blood samples at the time of presentation to hospital. Patients were categorized into asymptomatic (n=3), mild (n=7) and moderate (n=6) stages of COVID-19 severity. “Mild” disease is defined as the presence of COVID-19 disease in a patient who does not require hospitalization. “Moderate” disease is defined as the presence of COVID-19 disease in a patient who requires hospitalization. “Severe” disease is defined as the presence of COVID-19 disease in a patient who requires mechanical ventilation in an intensive care unit.

### Cohort 7

*<u>Singaporean cohort</u>*: The study was approved by the National Healthcare Group Domain Specific Review Board (DSRB 2014/00614). Written informed consent was obtained from all study participants. Study participants were individuals with suspected respiratory infections during the onset of the COVID-19 pandemic at the National University Hospital, Singapore. Individuals were considered COVID-19 patients if the patient tested positive for the virus by RT-PCR on admission (n=2). *<u>Australia cohort</u>*: This study has been approved by Research Governance at the Westmead Institute for Medical Research, the Human Research Ethics Committee at Western Sydney Local Health District (HREC Reference: 2020/ETH00886 (6439) and at Nepean Blue Mountain Local Health District (HREC Reference: 2019/ETH01485). Informed consent was obtained from all study participants. Study participants were individuals with suspected respiratory infections during the onset of the COVID-19 pandemic in the Southern Hemisphere (Westmead Hospital, Nepean Hospital, Sydney Australia and National University Hospital, Singapore). Subjects with recent (within the prior 14 days) vaccination history, infection/under antimicrobial medication, subjects under immunosuppressive drugs were not included in the study. Individuals became eligible for the study immediately upon the reporting of suspected COVID-19 symptoms (e.g., fever, sore throat, cough). Individuals were considered COVID-19 patients if the patient tested positive for the virus by qRT-PCR on admission or in the subsequent 28-day follow-up period.

### Cohort 8 – 10

The study has been approved by the Human Research Ethics Committee of Nepean and Blue Mountain Local Health District (HREC Reference: 11/26 – HREC/11/Nepean/46), Australia. Eligible patients included adult patients (>18 years) who presented with possible respiratory tract infection symptoms. To be eligible, the patient needed to have at least one symptom from two or more symptoms categories. The symptom categories are: (1) fever, (2) constitutional symptoms (chill, headache, muscle ache), (3) Respiratory symptoms (cough, sore throat, nasal congestion, or shortness of breath). After patient was enrolled into the study, venous blood was collected from the patient and placed into PAXgene. RNA extraction was performed as per manufacturer’s protocol. *IFI27* gene-expression was measured by quantitative real-time PCR. Researchers who performed PCR assays were blinded to information regarding patient data or etiological diagnoses. Nasopharyngeal, sputum, urine and blood samples were obtained at admission. These samples were sent for standard microbiological testing including sputum Gram stain and culture, blood culture. In addition, urinary antigen test for *Streptococcus pneumoniae* was also performed. In patients admitted to intensive care unit, additional respiratory samples were obtained from bronchoalveolar lavage or tracheal aspirates. In addition to standard microbiological tests, testing for atypical respiratory pathogens (*Chlamydophila pneumoniae, Mycoplasma pneumoniae* and *Legionella pneumonphila*) was also performed in selected patients at the discretion of treating physicians. SARS-Co-V2 virus was identified by virus PCR. In addition, respiratory viruses were tested in all patients using nucleic acid PCR in respiratory samples including sputum, nasopharyngeal or bronchoalveolar lavage (for patients in intensive care units). The PCR panel tested for influenza A, influenza B, respiratory syncytial virus, rhinovirus, parainfluenza virus and human metapneumovirus. All patients were followed up on study completion. Researchers retrieved relevant information from medical records using a pre-specified data collection form including outcomes, laboratory results, treatments, and microbiological reports. Telephone follow-up was performed where information was incomplete. Researchers who performed the follow-up were blinded to the *IFI27* levels.

### Experiments performed in Cohort 1

#### Rapid Autopsy Tissue

Formalin-fixed paraffin-embedded (FFPE) tissue blocks were prepared from autopsied pulmonary tissue from 4 COVID-19 patients who died from respiratory failure (ARDS). Autopsy and biopsy materials were obtained from the Pontificia Universidade Catolica do Parana PUCPR the National Commission for Research Ethics (CONEP) under ethics approval numbers 2020001792/30188020.7.1001.0020 and 2020001934/30822820.8.000.0020. The study was also approved under The University of Queensland Human Research Ethics Committee (HREC) ratification. Details of the patient cohort have been described previously [8].

### RNAscope^®^ of lung tissue

RNAscope^®^ probes (ACDbio, US) targeting SARS-CoV-2 spike mRNA (nCoV2019, #848561-C3) were used as per manufacturer instructions for automation on Leica Bond RX. DNA was visualised with Syto13 (Thermofisher Scientific. Fluorescent images were acquired with Nanostring Mars prototype DSP at 20x. RNAscope data from NanoString was analysed by the STRISH program (Tran et al, 2020). STRISH first performed cell segmentation and followed by mapping the SARS-CoV-2-positive cells based on RNAscope fluorescent signal (nCoV2019 spike mRNA). Subsequently, the software scanned through the whole tissue and counted cells with positive SARS-CoV-2 marker within segmented cells. STRISH gradually split the image into smaller windows until each window contains fewer than 100 cells. STRISH then performed min-max normalisation and plotted a heatmap to display the level of expression signal. The analysis code is available at the following weblink: https://github.com/BiomedicalMachineLearning/Covid19.

### Spatial transcriptomics (Visium)

Method 1 (PolyA-capture): FFPE tissue blocks were sectioned at 7μm thickness using a microtome and the section was transferred to a water bath at 41°C. The floating section was adhered to the Visium Spatial Gene Expression Slide (10x Genomics, USA, PN 2000233), and stored overnight at 4°C. The slide was deparaffinised by placing it on the surface of a thermomixer at 60°C for 30 minutes, immersing in Xylene for 5 mins twice and in ethanol six times, 2 minutes each. The H&E staining was carried out as per the Visium instructions, except that 100 μl of glycerol 85% was used. Tissue imaging was performed using an Axio Scan Z1 Fluorescent Slide Scanner. After H&E imaging, the slide went through pre-permeabilisation (37°C, with collagenase mix for 20 mins), decrosslinking (70°C for 1 hour, in TE buffer pH 8.0) and permeabilization (75 μl 0.1% Pepsin, 37°C for 10 minutes). Library preparation was performed according to the Visium Gene Expression user guide (CG000239).

Method 2 (Probe-based hybridization): FFPE tissue blocks were prepared, sectioned, and placed on Visium Spatial Gene Expression Slides according to the tissue preparation protocol (CG000408). Subsequent processing, permeabilization, and library preparation were performed according to the Visium Spatial Gene Expression for FFPE user guide (CG000407).

The Visium raw sequencing data in BCL format was converted to 137,604,799 FASTQ reads using bcl2fastq/2.17. The reads were trimmed by cutadapt/1.8.3 to remove poly-A tails and template-switching-oligos (slide V10N16-049-B1). We used SpaceRanger V1.2.2 to map FASTQ reads to the CellRanger human reference genome and gene annotation for GRCh38-3.0.0. On average, for each spot we mapped 45,914 reads and detected a total of 17,598 genes, with an average of 565 genes per spot. We applied an adaptive binning strategy for the visualisation of gene expression for individual spots across all tissues. Four bins were calculated: off, low, medium, and high expression. The low to high bins were defined separately for each gene based on equal division of their normalised expression; genes with no counts were added to the off bin. Differentially expressed genes between virus-high and virus-low samples were identified using a negative binomial test as implemented in Seurat v4 (25).

### ELISA performed in Cohorts 4 & 5

A volume of 4.0ml of whole blood was collected in EDTA tubes (BD Frankilin Lakes, NJ, USA). The tubes were centrifuged for 10 minutes at 400g, and the plasma obtained was stored at -80°C for long term storage. 25ul of plasma from these patients was analysed in duplicate using IFI-27 ELISA kit (Aviva Systems Biology, USA). Serum IFI-27 (pg/ml) were log2 transformed and median centred to evaluate the relationship with clinical annotation.

## SUPPLMENTARY FIGURE

Three representative patients (A, B and C) from Cohort 7 are presented here:

1. The clinical trajectory of each patient is plotted along a time-axis (in days), which is located at the top of the graph. Along this axis, the day of hospital admission or ICU admission is indicated by green colour.
2. Within each graph, a plus or minus sign (“+” or “-”) indicates the presence or absence of an observation (e.g. fever) or a measurement (e.g. positive SARS-CoV-2 virus). If an observation or measurement was not performed, then an “o” sign is provided.
3. A colour scale (located on the right-hand side of the graph) is used to indicate the degree of abnormality in each observation or measurement. Blue colour indicates the observation or measurement is within the normal range. Light red colour indicates a mildly abnormal observation or measurement. Deep red colour indicates a highly abnormal observation or measurement.
4. The *IFI27* gene expression is expressed as fold change.
5. The **Supplementary Figure** is shown on the next page.

### Patient A

#### History and physical examination

This patient presented with 7 days of fever and cough. On the day of hospital admission, the patient had a mildly elevated heart rate (101 – 120/min), but other vital signs were normal, including respiratory rate (< 20 breath/min) and blood pressure (mean systolic pressure 66 – 100 mmHg). In keeping with the clinical history (suspected COVID-19), the patient had a moderately elevated temperature (39 – 40 degree Celsius) on admission. At the time of physical examination, the patient was not distressed and did not require any supplemental oxygen therapy (inspired oxygen concentration was normal at 21%).

#### Laboratory findings

The patient was test positive for SARS-CoV-2. The leucocyte count on admission was normal, although the neutrophil-lymphocyte ratio was mildly abnormal (4.5). Otherwise, there was no other abnormal laboratory findings.

#### IFI27 level

The *IFI27* level on presentation was abnormal and extremely high (**707**).

#### Clinical course

The patient began to show signs of hypoxaemia on the second day after admission, with an increased respiratory rate (21–30 breath/min) and required supplemental oxygen (inspired oxygen concentration at 22%-29%). A further deterioration occurred on the third day where his inspired oxygen requirement increased to 30% - 49%. On the fourth day, he developed hypoxic respiratory failure and was admitted to intensive care unit for mechanical ventilation.

#### Summary

Abnormal *IFI27* biomarker level precedes signs of respiratory failure by at least 24 hours.

**Supplementary Figure.**
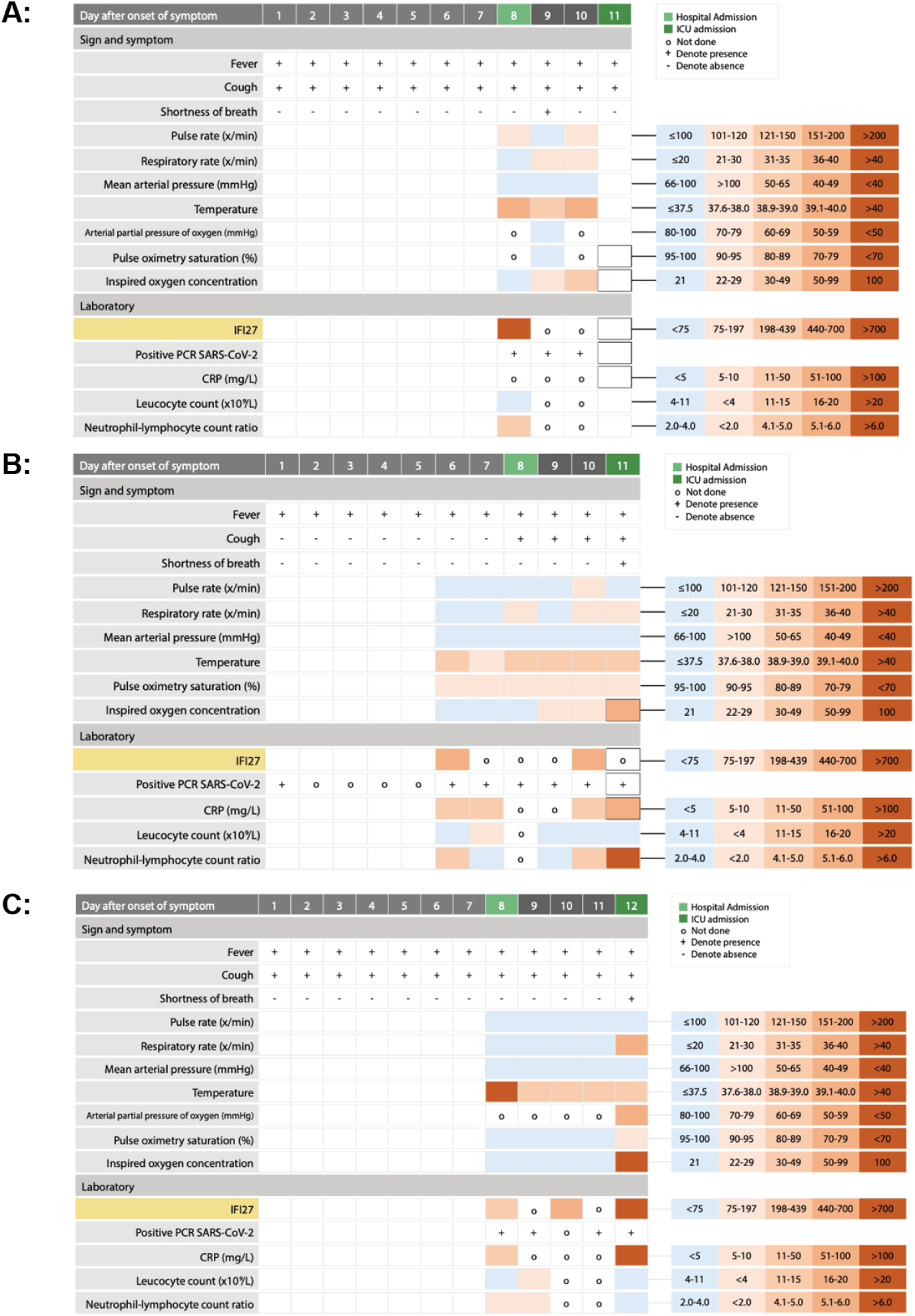

### Patient B

#### History and physical examination

This patient had no abnormal signs or symptoms on presentation, except for the presence of a fever (temperature 38.0 – 39.0 degree Celsius). The pulse oximetry saturation reading was a bit on the low side; but it was still within the normal range (90 – 95% saturation while breathing at room air).

#### Laboratory findings

The patient was test positive for SARS-CoV-2. Laboratory findings showed mildly elevated C-reactive protein (CRP) levels on day 6 (CRP: 12) and on day 7 (CRP: 22), and neutrophil-lymphocyte ratio (4.5). At the time, these findings did not warrant any concern.

#### IFI27 level

However, the *IFI27* level on the same day was highly abnormal (**446**).

#### Clinical course

The patient become tachypnoeic two days later, with respiratory rate increased to 21 – 30 breath/min. He was admitted to hospital and given supplementary oxygen therapy. On the fourth day after admission, he developed hypoxic respiratory failure and was admitted to intensive care unit for mechanical ventilation. It was noted, on the day of admission to the intensive care unit, his neutrophil-lymphocyte ratio rose sharply to a highly abnormal level (6.8). A repeated *IFI27* on the same day also showed an abnormal result (**459**).

#### Summary

This *IFI27* warning sign preceded changes in bedside measurement or laboratory findings by several days. The *IFI27* level was already abnormally elevated prior to admission to intensive care unit - indicating the patient had an early sign of disease progression, despite the fact the vital signs and other bedside parameters were relatively “normal” at the time.

### Patient C

#### History and physical examination

This patient presented with a high fever and cough. At the time of assessment, he had no abnormal physical signs and had no evidence of hypoxaemia.

#### Laboratory findings

The patient was test positive for SARS-CoV-2. Laboratory findings showed a mildly elevated C-reactive protein level (**16**), which did not warrant any concern.

#### IFI27 level

The *IFI27* level on the admission day was moderately abnormal (**226**).

#### Clinical course

The patient was admitted to hospital for monitoring because of positive SARS-CoV-2 status. The patient remained stable for the following three days and showed no signs of distress or hypoxaemia. However, a repeated *IFI27* measurement on the second day after admission revealed a rapidly rising *IFI27* level (**672**), which suggested an increasing risk of deterioration. On the fourth day, the patient suddenly developed severe hypoxic respiratory failure which required urgent admission to intensive care unit. On the same day, a further rise in *IFI27* level was noted (**881**), as was a highly abnormal C-reactive protein level (247 mg/L). Interestingly, on the day of deterioration, both leukocyte count and neutrophil-lymphocyte ratio remained within the normal range.

#### Summary

In this patient, the rise in *IFI27* levels occurred prior to clinical deterioration. Again, laboratory findings and changes in vital signs lagged behind *IFI27* rise.

**SUPPLEMENTARY TABLE 1.**

| **COVID-19 related symptoms** | **ISARIC-defined COVID-19 complications** |
| --- | --- |
| Fever | Viral pneumonia |
| Cough | Bacterial pneumonia |
| Sore throat | ARDS |
| Runny nose | Cardiac complications |
| Myalgia | Bacteraemia |
| Arthralgia | DIC |
| Fatigue | Liver dysfunction |
| Chest pain | Stroke/CVA |
| Shortness of breath | Seizure |
| Headache | Acute kidney injury |
| GIT symptoms (e.g., diarrhoea) | Hyperglycaemia |
| Loss of smell | Pulmonary embolism |
| Loss of taste | Deep venous thrombosis |

ISARIC denotes International Severe Acute Respiratory and Emerging Infection Consortium.

**SUPPLEMENTARY TABLE 2**

This is attached as an Excel spreadsheet titled “Supplementary Table 2.xls”

